# Covid-19 Pandemic and its Effect on Residents’ Mental Well-Being

**DOI:** 10.1101/2021.11.12.21266292

**Authors:** Anwar Yazdani, Hend Esmaeili, Abdulla K AlSaleh, Ahmed Sultan, Esam Alamad, Ali Bandar, Hanouf Rawdhan, Mariam Ayed

**Author notes:** **Corresponding author:** Mariam Ayed, Neonatal Department, Farwaniya Hospital, Subah An Nasser, Kuwait City, Postal code-81400, Kuwait, E mail. **Funding:** None.

## Abstract

Concerns about COVID-19’s long-term consequences on the mental health of frontline health professionals are mounting as the entire world strives anew to contain it. The primary objective of this research is to describe the impact of working during the COVID-19 pandemic on junior doctors’ mental health and to investigate the effect of the COVID-19 pandemic on junior doctors’ training and professional performance. A cross-sectional online survey using the Google Forms platform was conducted from May 1^st^ to May 30^th^, 2021, in 311 healthcare workers who were currently enrolled in a residency program at the Kuwait Institutional of Medical Specialization (KIMS). Socio-demographic details of each health worker were collected and the scores related to depression, anxiety, and stress were measured using the previously validated depression anxiety stress scale-21 (DASS-21). Higher stress scores were seen in those who were devoid of the option to work with COVID-19 patients (adjusted β 5.1 (95%CI:1.2-9);*p=*0.01), who reported that working during the pandemic affected their study schedule (adjusted β 4.8 (95%CI:1.6-8.1);*p=* 0.004), and who lost off service training time (adjusted β 2.7 (95%CI:0.13-5.2); *p=*0.034). Further, the anxiety scores were significantly higher in females. The impact of the ongoing pandemic on residents’ mental health is grave, necessitating psychological treatment and support. The study discovered various factors linked to depression, anxiety, and stress. As a result, these aspects must be regarded to protect the residents’ mental health.

## Introduction

The Covid-19 pandemic evolved from an immediate health emergency to a systemic problem that had far-reaching consequences in people’s lives. Because of its progression from a health scare to a global economic and social disaster, the effects of Covid-19 have been unparalleled (Conceição et al., 2020).

Past investigations of the effects of the Severe Acute Respiratory Syndrome, Middle East Respiratory Syndrome, and Ebola epidemics on suffering individuals and healthcare workers revealed a significant neuropsychiatric connection (Shah et al., 2020). Covid-19’s effects on people’s mental health and well-being were being explored more and more (Torales et al., 2020; Tang et al., 2020). Even prior to the COVID-19 pandemic, a considerable body of research has shown that resident physicians experience burnout, depression, and anxiety while in training (Raj, 2016). As a result, the possibility of a reduction in their well-being due to the COVID-19 pandemic is very plausible.

Kuwait’s national healthcare system, like that of many other countries, was also strained during the COVID-19 epidemic (Armocida et al., 2020). Even in high-income countries with well-resourced health care systems, overburdened healthcare services confronted considerable operational and logistical issues. As a result, many healthcare staff have been deployed due to the pandemic response (Coughlan et al., 2020). Many junior doctors, despite their specializations, were associated with the management of COVID-19 patients in a variety of setups such as quarantine areas, swabbing areas, emergency departments, COVID-19 wards, and ICUs, starting from February 24^th^ (when the first case of COVID-19 was diagnosed in Kuwait) to the end of September 2020. These kinds of scenarios were also documented in a number of other nations. Junior doctor rotations were reported to be discontinued in the United Kingdom (UK), and many doctors were redeployed to operate in intensive care units (Salem et al., 2020). The specific approach to tackle the epidemic incorporates a number of interconnected ideas, and multiple studies have found that capability outcomes are closely linked to mental health and social consequences (Brunner, 2017). Despite the growing number of studies looking into the influence of COVID-19 on mental health and well-being, data on the pandemic’s broader potential impact is still lacking. Hence, this study aimed to recognize the participation of junior doctors in Kuwait in the COVID-19 pandemic and note their ability to deal with the tragedy and its impact on their mental health and professional training.

The major objective of the present study includes describing the effect of working during the pandemic on the mental well-being of junior doctors in Kuwait as well as to study the effect of the COVID-19 pandemic on the training and professional performance of the junior doctors in Kuwait.

## Method

A cross-sectional online survey was created to assess mental and physical health as a result of COVID-19 exposure. The Google Forms platform was used to conduct this survey from 1^st^ May to 30^th^ May 2021. Health-care workers who were currently enrolled in a residency program at the Kuwait Institutional of Medical Specialization (KIMS) were eligible to participate in this study. The study was authorized by Kuwait’s Ministry of Health’s Ethical Research Committee. The participants gave their consent to participate in the survey by filling the questionnaire.

Each residency program’s director, chief residents, and head of medical departments received the questionnaire by email or WhatsApp. The participants were then requested to forward the survey invitation to other junior doctors, with a two-week follow-up reminder. Limited response to one individual was enabled in the Google forms to avoid multiple submissions from the same participant. The information gathered was automatically placed into a spreadsheet.

### Data collection

The sociodemographic details such as age, gender, marital status, and nationality of each patient were collected. In addition, the physicians were questioned about their participation during the pandemic, whether they had the choice to work with COVID-19 patients, whether they had mentor or supervisor assistance, and whether they had received sufficient personal protective equipment (PPE) training.

The dependent variables such as depression, anxiety, and stress were measured using the previously validated depression anxiety stress scale-21 (DASS-21) (Lovibond & Lovibond, 1995). The DASS-21 is divided into the following subscales, each with seven items: depression, anxiety, and stress. A 4-point Likert scale is used in the DASS. The possibilities for responses vary from “Never” to “Almost Always” (0–3). The total of all subscale elements determines the subscale score. The cut-offs for the stated divisions can be enumerated as: Depression (normal/mild = 0–13; moderate = 14–20; severe/extremely severe = 21+), anxiety (normal/mild = 0–9; moderate = 10–14; severe/extremely severe = 15+), and stress (normal/mild = 0–18; moderate = 19–25; severe/extremely severe = 26+).

### Statistical analysis

STATA 14 IC was used to analyze the data. Descriptive statistics were expressed as a number and a percentage for categorical variables, while for continuous data, the median and interquartile range (IQR) were used. The DASS-21 scores were compared to the demographic and survey responses of the individuals using the Mann Whitney U test or the Kruskal Wallis test. The multiple linear regression included all variables with a P-value of less than 0.05 in the univariate analysis. The significant risk factors related to stress, depression, and anxiety were confirmed using backward stepwise linear regression analysis. Variance inflation factor (VIF) statistics were used to test multicollinearity. The analysis was run in separate models to see how the variables, depression, anxiety, and stress (DASS-21), behaved separately in the multivariate model as they were noted to be multicollinear. In regression analysis, the strength of the link was expressed as a beta (ß) coefficient and a 95 % confidence range (95% CI). All tests were two-tailed, with an alpha of 0.05.

## Result

### Participants’ characteristics

A total of 311 responders completed the survey and were included in the present study. Around 50.8% of the participants were single, 50.2% were female, and 58% were in the age group of 25-29 years. The socio-demographic details of the enrolled participants are presented in Table 1.

**Table 1.**
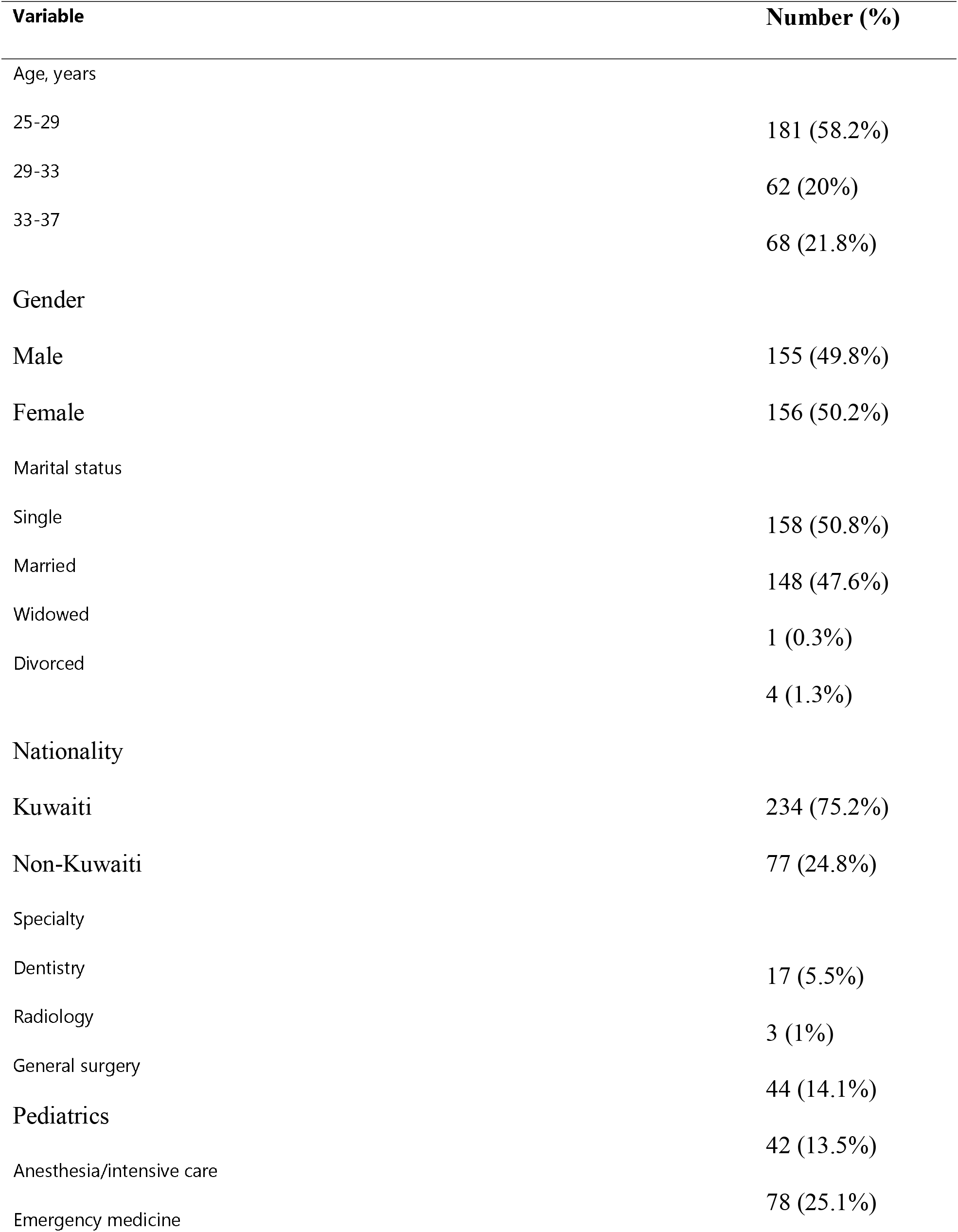
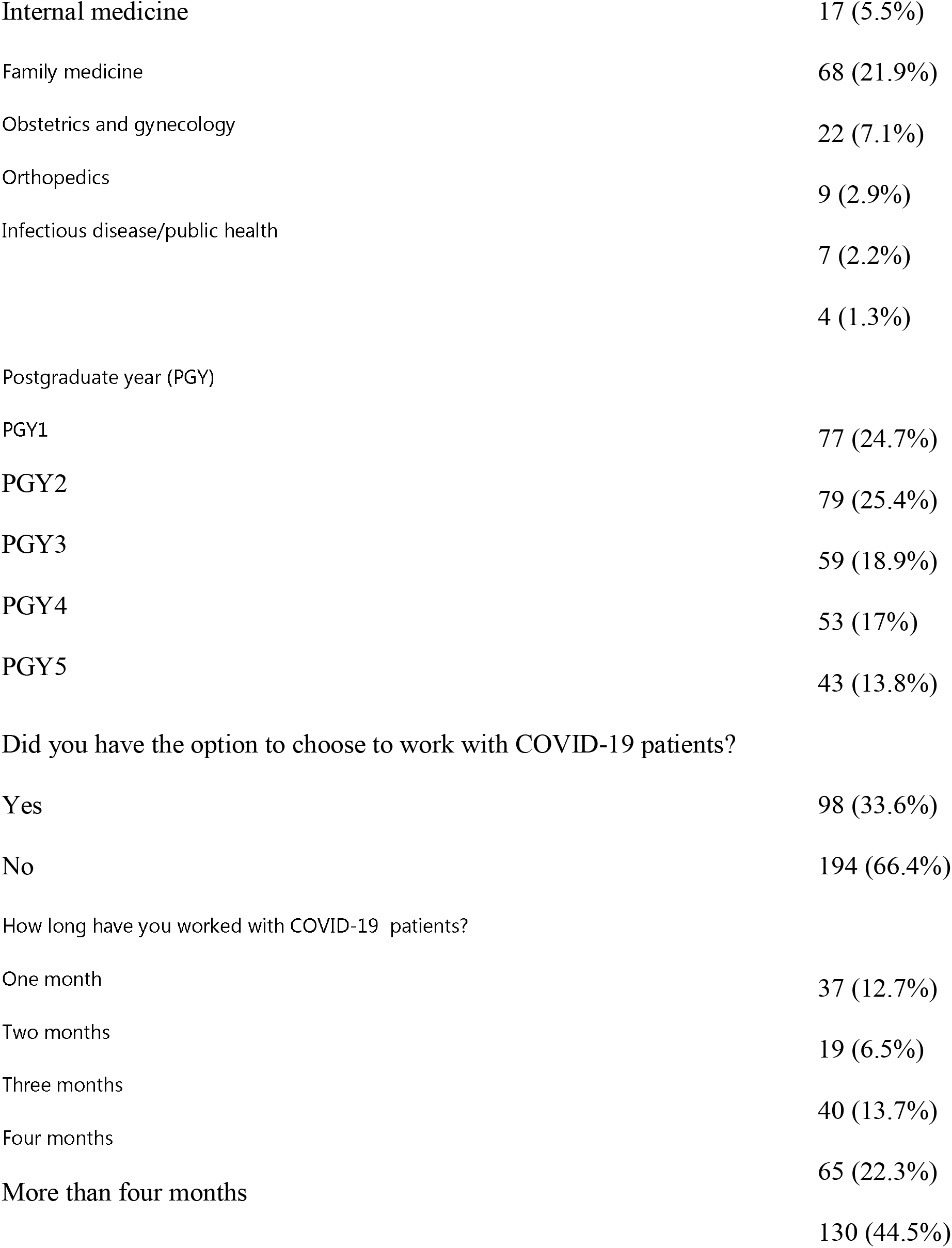
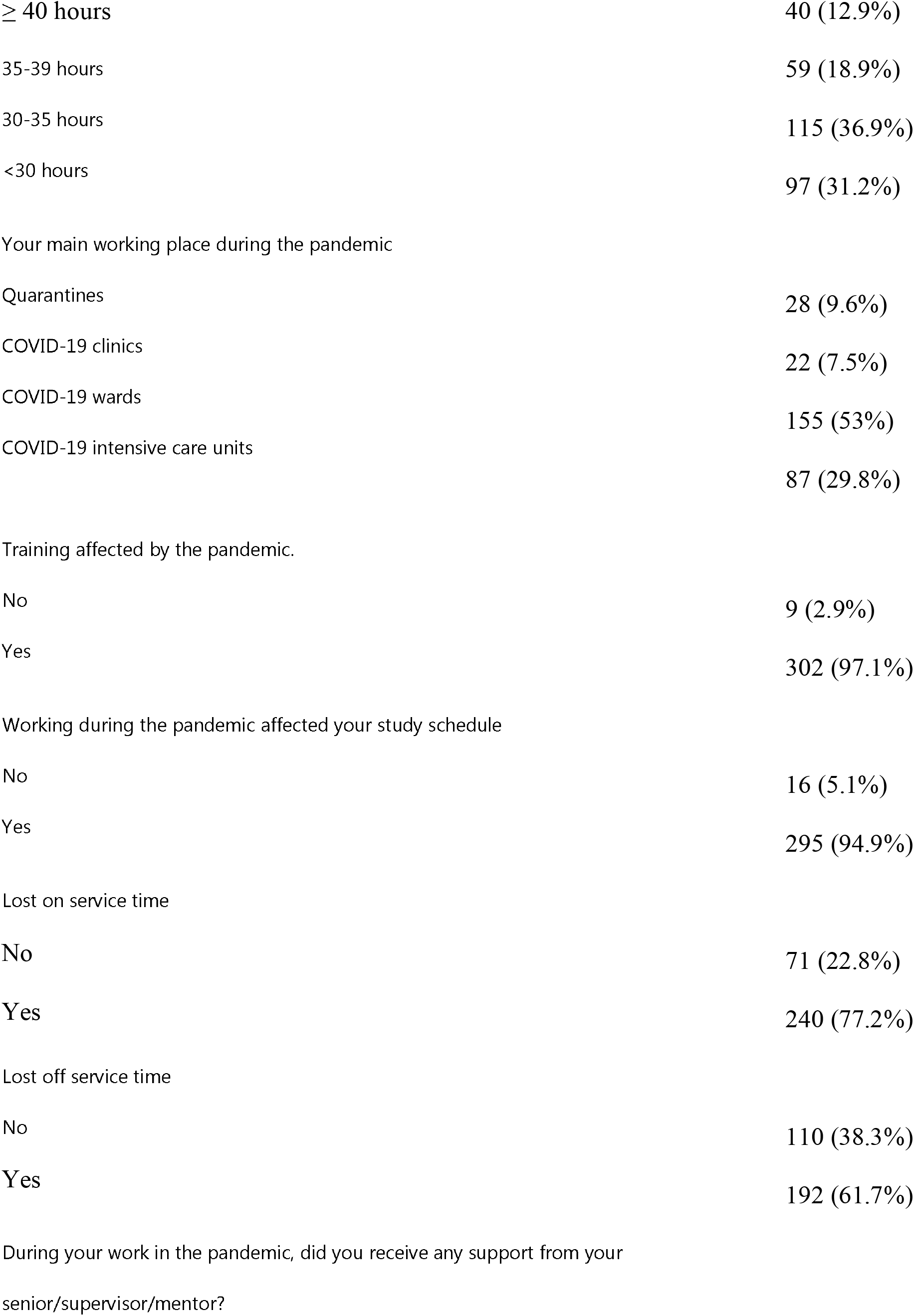
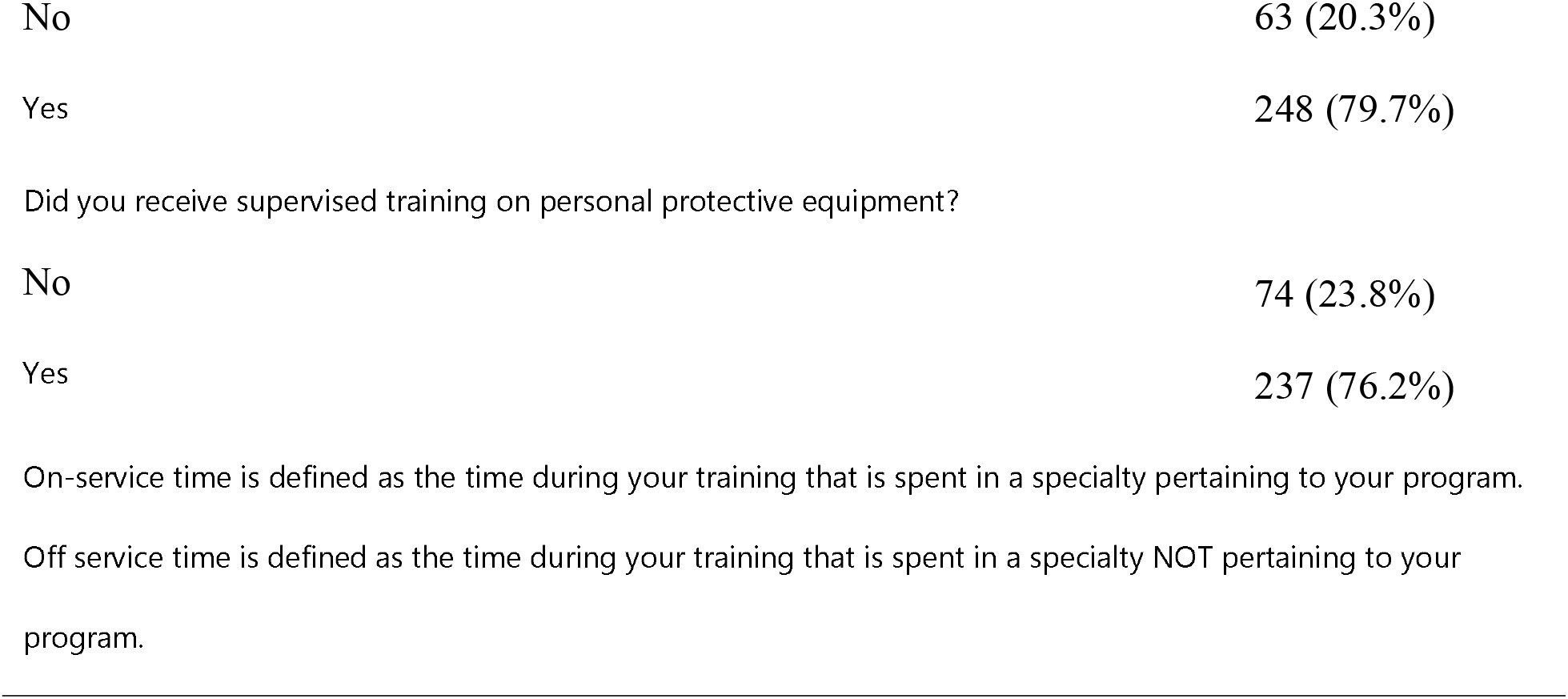
Sociodemographic characteristics of the participants

| Variable | Number (%) |
| --- | --- |
| Age, years |  |
| 25-29 | 181 (58.2%) |
| 29-33 | 62 (20%) |
| 33-37 | 68 (21.8%) |
| Gender |  |
| Male | 155 (49.8%) |
| Female | 156 (50.2%) |
| Marital status |  |
| Single | 158 (50.8%) |
| Married | 148 (47.6%) |
| Widowed | 1 (0.3%) |
| Divorced | 4 (1.3%) |
| Nationality |  |
| Kuwaiti | 234 (75.2%) |
| Non-Kuwaiti | 77 (24.8%) |
| Specialty |  |
| Dentistry | 17 (5.5%) |
| Radiology | 3 (1%) |
| General surgery | 44 (14.1%) |
| Pediatrics | 42 (13.5%) |
| Anesthesia/intensive care | 78 (25.1%) |
| Emergency medicine |  |
| **Internal medicine** | 17 (5.5%) |
| Family medicine | 68 (21.9%) |
| Obstetrics and gynecology | 22 (7.1%) |
| Orthopedics | 9 (2.9%) |
| Infectious disease/public health | 7 (2.2%) |
|  | 4 (1.3%) |
| Postgraduate year (PGY) |  |
| PGY1 | 77 (24.7%) |
| **PGY2** | 79 (25.4%) |
| PGY3 | 59 (18.9%) |
| PGY4 | 53 (17%) |
| PGY5 | 43 (13.8%) |
| **Did you have the option to choose to work with COVID-19 patients?** |  |
| Yes | 98 (33.6%) |
| No | 194 (66.4%) |
| **How long have you worked with COVID-19 patients?** |  |
| One month | 37 (12.7%) |
| Two months | 19 (6.5%) |
| Three months | 40 (13.7%) |
| Four months | 65 (22.3%) |
| **More than four months** | 130 (44.5%) |
| Work schedule |  |
| $\geq 40$ hours | 40 (12.9%) |
| 35-39 hours | 59 (18.9%) |
| 30-35 hours | 115 (36.9%) |
| <30 hours | 97 (31.2%) |
| Your main working place during the pandemic |  |
| Quarantines | 28 (9.6%) |
| COVID-19 clinics | 22 (7.5%) |
| COVID-19 wards | 155 (53%) |
| COVID-19 intensive care units | 87 (29.8%) |
| Training affected by the pandemic. |  |
| No | 9 (2.9%) |
| Yes | 302 (97.1%) |
| Working during the pandemic affected your study schedule |  |
| No | 16 (5.1%) |
| Yes | 295 (94.9%) |
| Lost on service time |  |
| No | 71 (22.8%) |
| Yes | 240 (77.2%) |
| Lost off service time |  |
| No | 110 (38.3%) |
| Yes | 192 (61.7%) |
| During your work in the pandemic, did you receive any support from your senior/supervisor/mentor? |  |
| No | 63 (20.3%) |
| Yes | 248 (79.7%) |
| Did you receive supervised training on personal protective equipment? |  |
| No | 74 (23.8%) |
| Yes | 237 (76.2%) |

On-service time is defined as the time during your training that is spent in a specialty pertaining to your program.
Off service time is defined as the time during your training that is spent in a specialty NOT pertaining to your program.

One hundred and ninety-four (66.4%) participants reported that they lacked the option to choose to work with COVID-19 patients. Most of KIMS residents (97.1% and 94.9% believed that their training and study schedule were affected by the pandemic, respectively. Lost on and off service training time was reported in 77.2% and 61.7% of the residents, respectively. The majority of the registered participants (79.7%) indicated they were devoid of getting support from their supervisors/mentors, and 76.2 % of participants stated they lacked adequate PPE training.

### DASS-21 results

The median DASS-21 score was noted to be 16 (IQR: 10-24) for stress, 12 (IQR:6-24) for depression, and 10 (IQR: 4-18) for anxiety. The observed values inferred the fact that residents had moderate symptoms of depression and anxiety and normal/mild symptoms of stress.

Table 2 shows the median (IQR) scores of stress, depression, and anxiety stratified by the participants’ characteristics and responses.

**Table 2.**
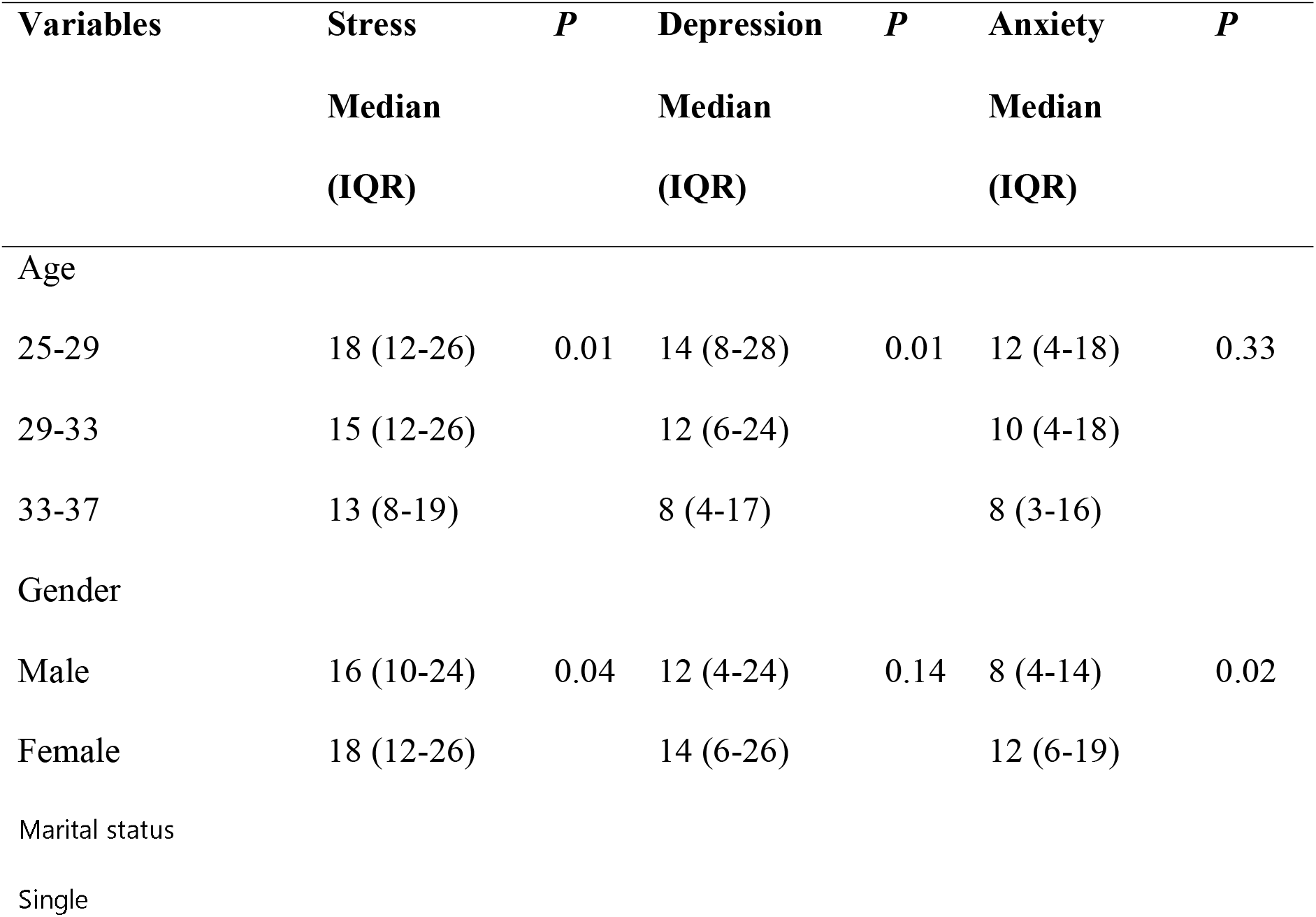
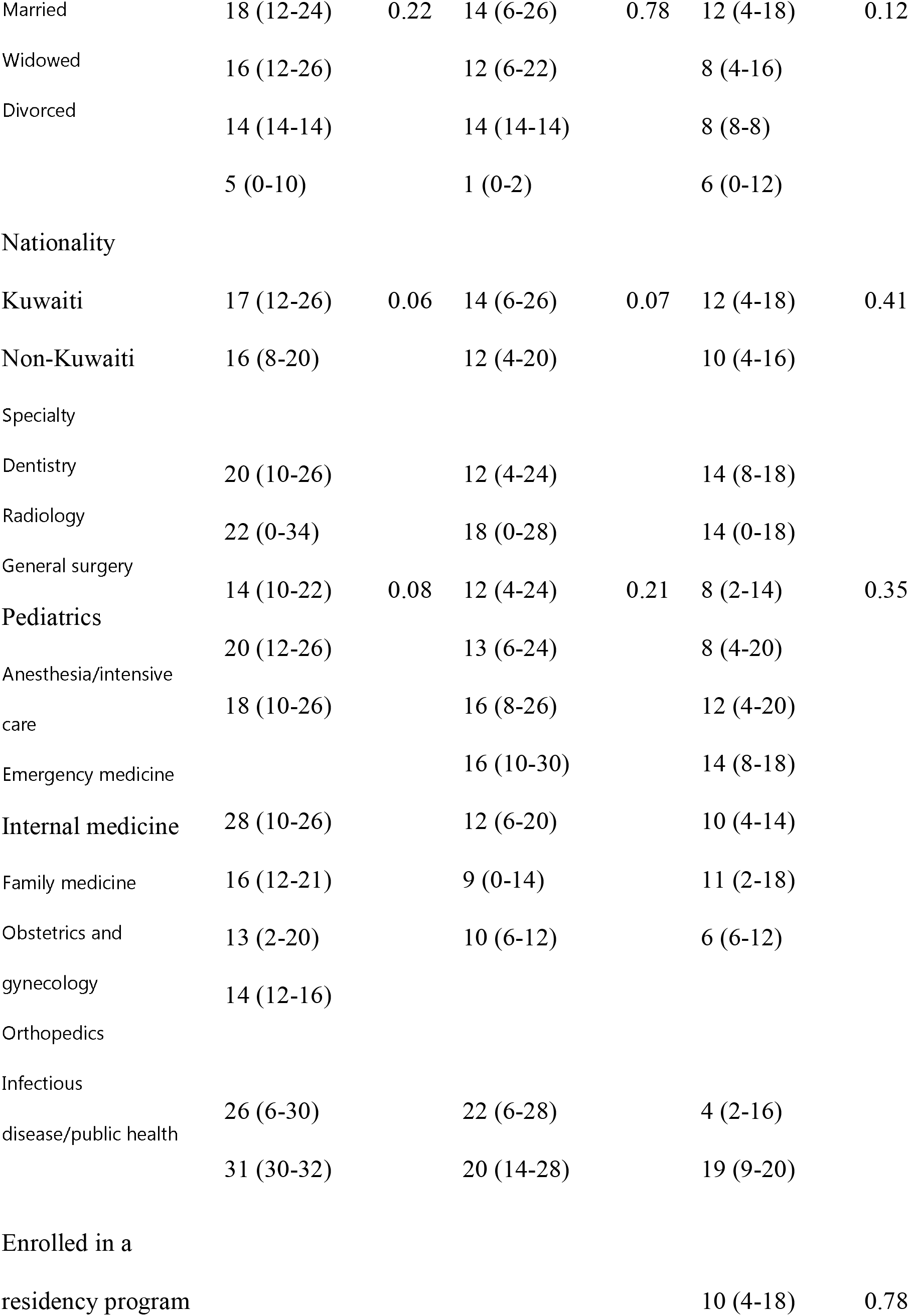
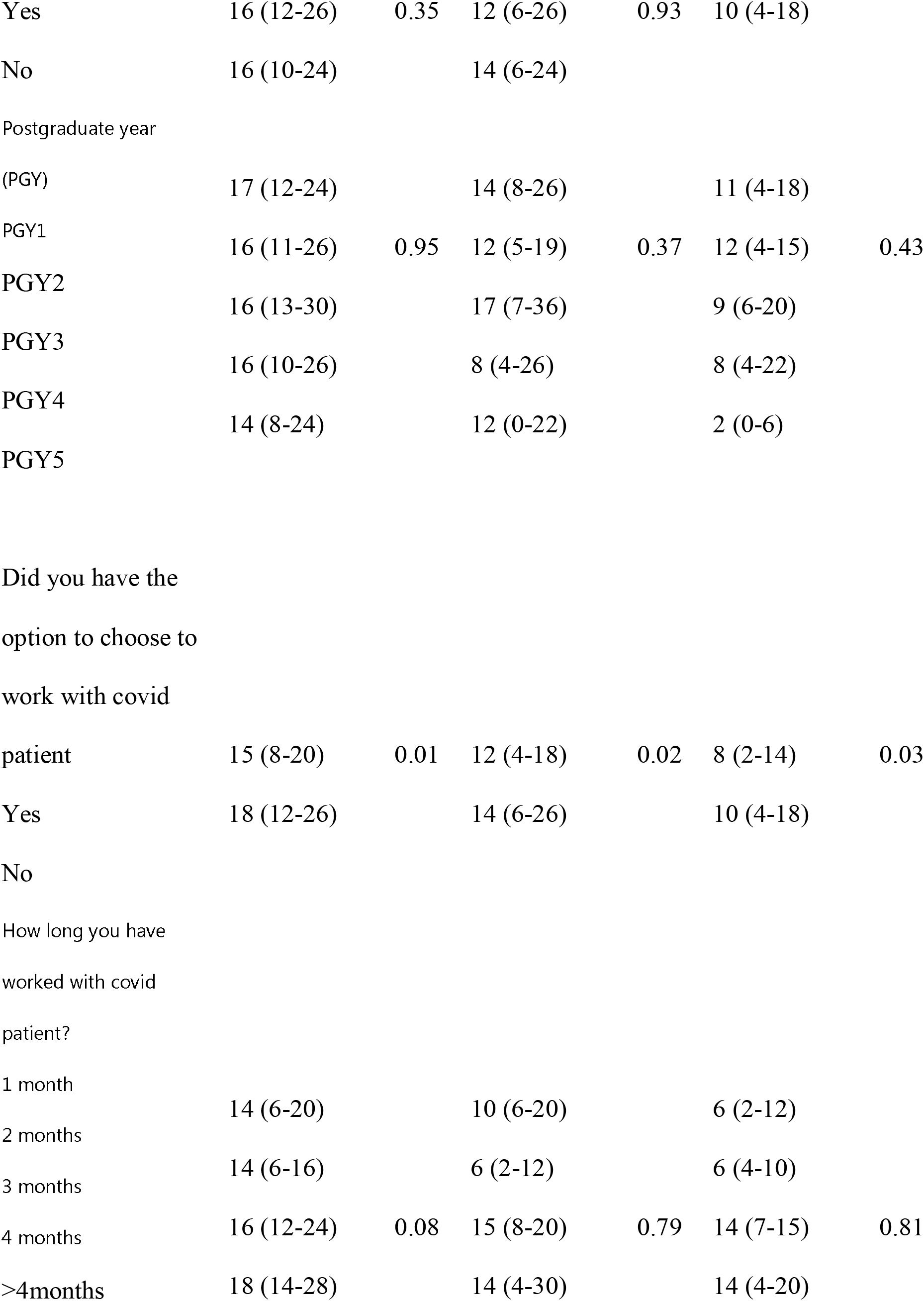
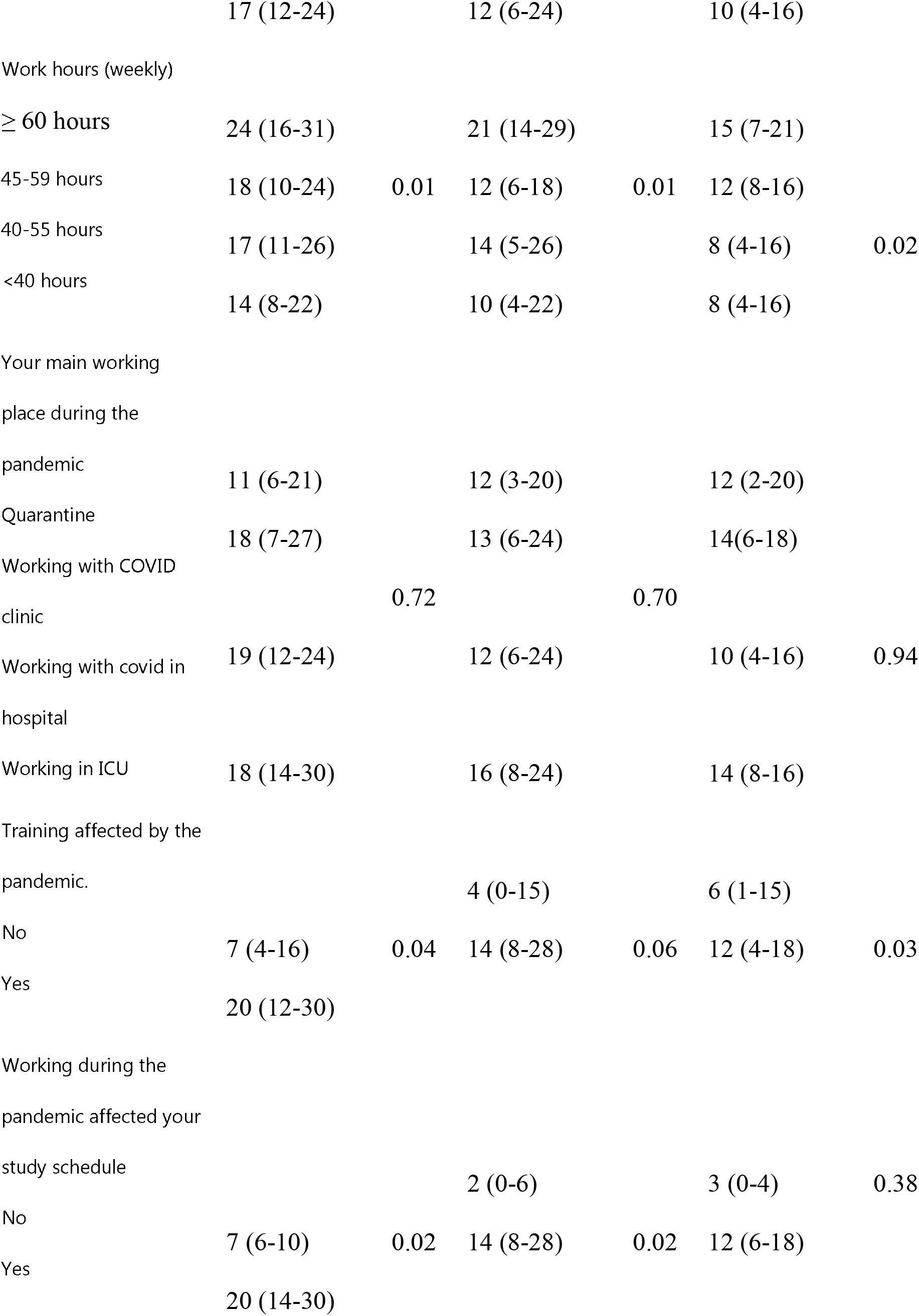
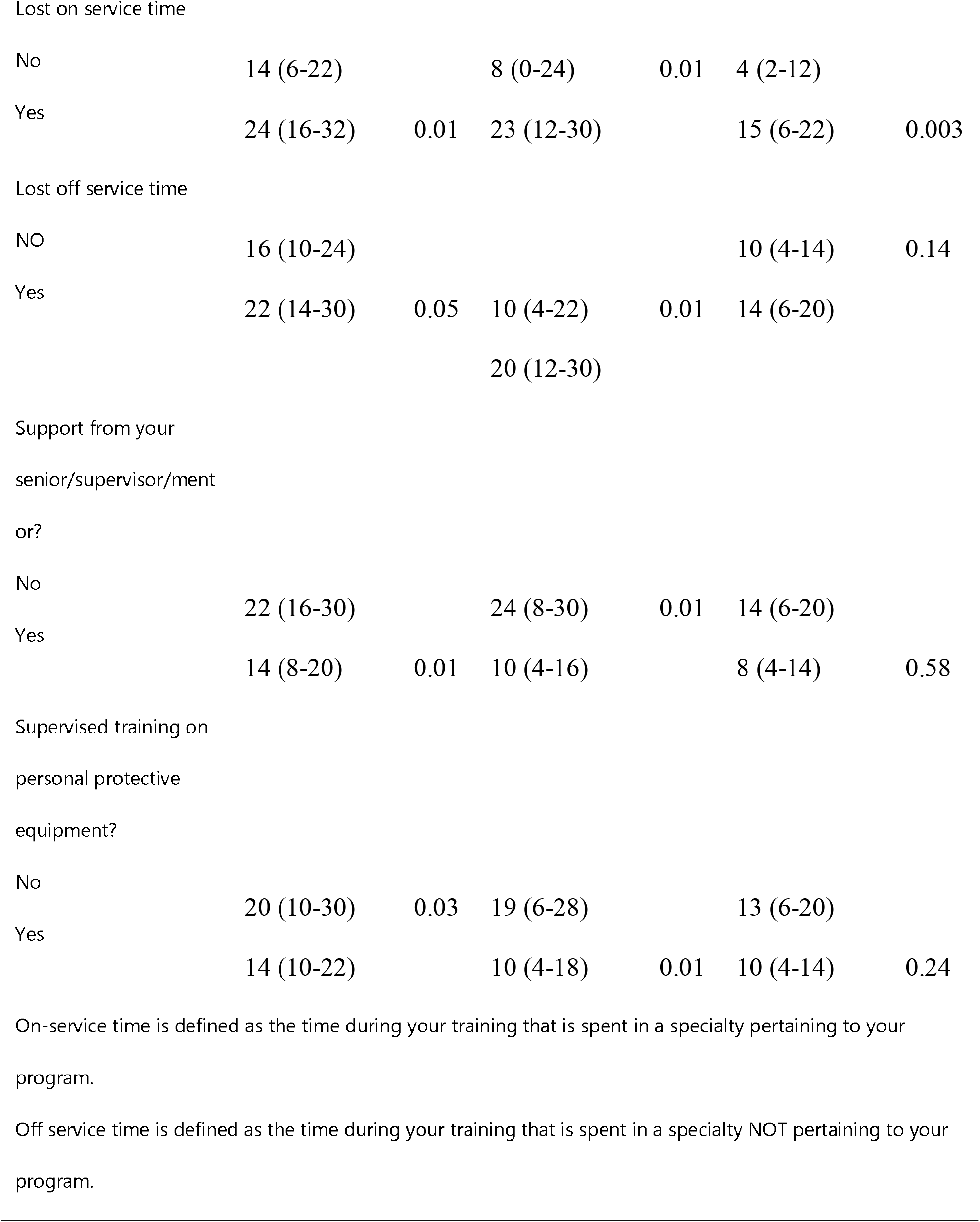
Analysis of DASS-21 score based on demographic and participants response

### Stress score

DASS-21 scores revealed that the prevalence of mild, moderate, severe, and extremely severe stress among the respondents was 14.6%, 17.5%, 13.7%, and 10.2%, respectively. The scores of stress were significantly higher among those aged 25-29 years compared to those aged 33-37 p<0.005). The stress score was significantly higher in females compared to males (p=0.04). Respondents who believed that their training and study schedule were affected by the pandemic, who lost on service training time, and who did not have the option to choose to work with COVID-19 patients had significantly higher stress scores. Moreover, those who had weekly work hours of more than 60 hours and those who did not receive mentor/supervisor support and proper PPE training had significantly higher stress scores.

### Depression score

The DASS 21 scores inferred that 12.7%, 19.4%, 10.2%, and 20.1% of the respondents were observed to have mild, moderate, severe, and extremely severe depression. The score for depression was significantly higher among those aged 25-29 years than those aged between 29-33 years(p=0.01). Participants who lost on and off service training time had a higher depression score than those who did not lose on and off service training time (p= 0.01). Further, respondents who had weekly work hours of more than 60 hours (p=0.01) had higher median depression scores. Depression scores were significantly higher in those who had no mentor or supervisor support (p=0.01) and proper PPE training (p=0.01) than those who had mentor or supervisor support and proper PPE training.

### Anxiety score

The proportion of mild, moderate, severe, and extremely severe anxiety was 13.7%, 17.8%, 8.6%, and 21.7%, respectively in the studied population.

The anxiety score was significantly higher among females compared to males (p=0.02). In addition, those who lost on service training time (p=0.003), those who did not have the option to choose to work with COVID-19 patients (p=0.03), training affected by pandemic (p=0.03), and worked more than 60 hours/week (p=0.02) had higher median anxiety scores.

The variables considered for analyzing their effect on depression, anxiety, and stress subscale scores include age, gender, marital status, nationality, course specialty, current program year of enrolled participants, option to choose to work with COVID-19 patients, time period worked with COVID-19 patients, work schedule, main working place during the pandemic, the effect of training and working schedule during the pandemic, loss on and off training time, any support from your senior/supervisor/mentor and receiving supervised training on personal protective equipment.

On multivariate linear regression analysis, higher stress scores were seen in those who were devoid of the option to work with COVID-19 patients (β=5.1, 95%CI: 1.2-9; *p=*0.01), who reported that working during the pandemic affected their study schedule (β= 4.8, 95%CI:1.6-8.1; *p=* 0.004), and who lost off service training time (β=2.7, 95%CI: 0.13-5.2; *p=*0.034) (Table 3). Similarly, higher depression scores were observed in those who did not have the option to work with COVID-19 patients (β= 6.5, 95%CI: 2.1-10.8; *p=*0.004), who reported that working during the pandemic affected their study schedule (β= 5.8, 95%CI: 2.3-9.3; *p=* 0.001), and who lost off service training time (β= 3.2, 95%CI: 0.69-5.7; *p=*0.013) (Table 4). Further, anxiety scores were significantly higher in females (β=3.1, 95%CI: 0.09-5.2; *p=*0.04), among those who feel that their training has been affected by the pandemic (β=2.9, 95%CI: 0.04-5.9; *p=*0.03) and who lost on service training time (β= 2.5, 95%CI: 0.11-4.8;*p=*0.04) (Table 5).

**Table 3.** Regression analysis for stress

| **Variable** | **Beta coefficient (95% confidence interval);**<br>**P value** |
| --- | --- |
| Doctors who didn't have the option to choose to work with COVID patient | 5.1 (1.2-9); 0.01 |
| Doctors feel that working during pandemic affected their study schedule | 4.8 (1.6-8.1); 0.004 |
| Doctors feel that their training has been affected by the pandemic | 2.8 (-0.56-6.2); 0.101 |
| Lost off service | 2.7 (0.13-5.2); 0.034 |
| Adjusted for age, gender, option to work with COVID-19 patients, working for more than 45 hours/week, training affected by the pandemic, study schedule affected by the pandemic, lost on service, lost off service, mentor support, and proper training on personal protective equipment. |  |

**Table 4.**
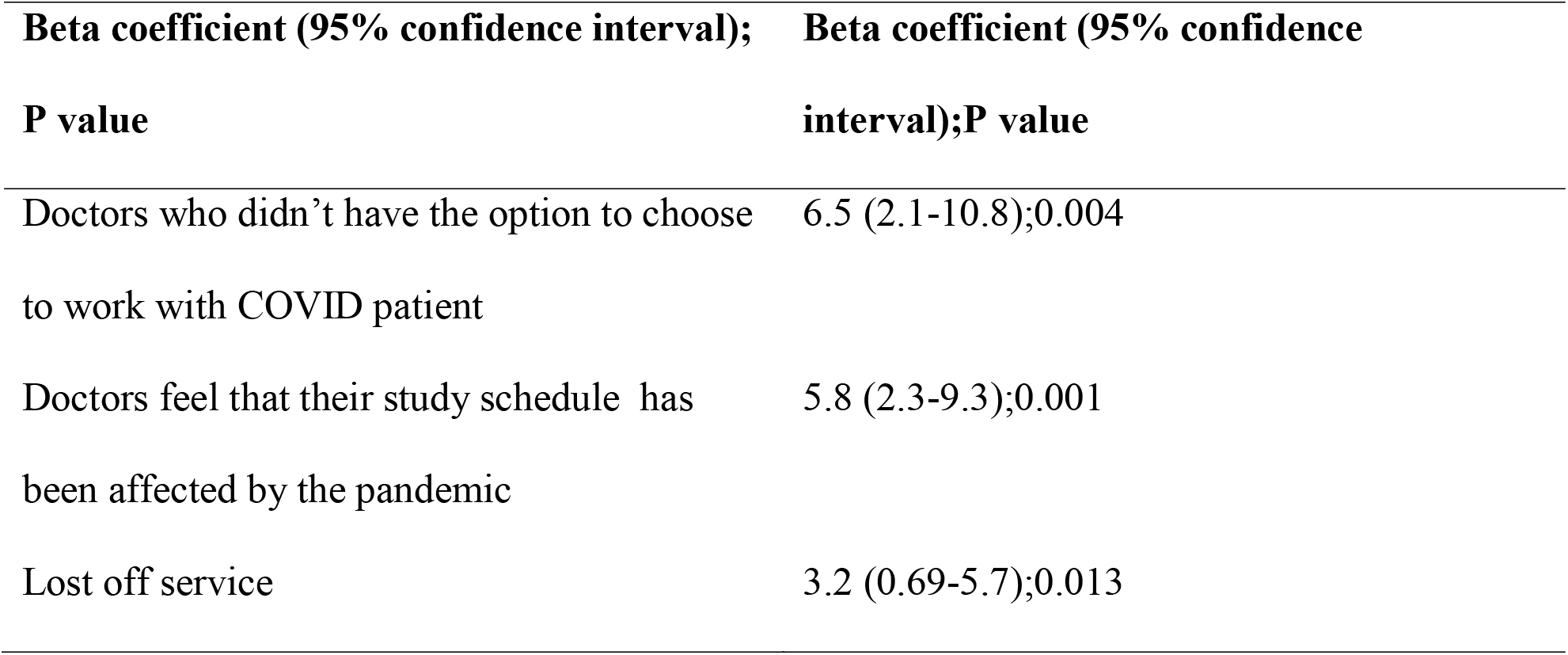
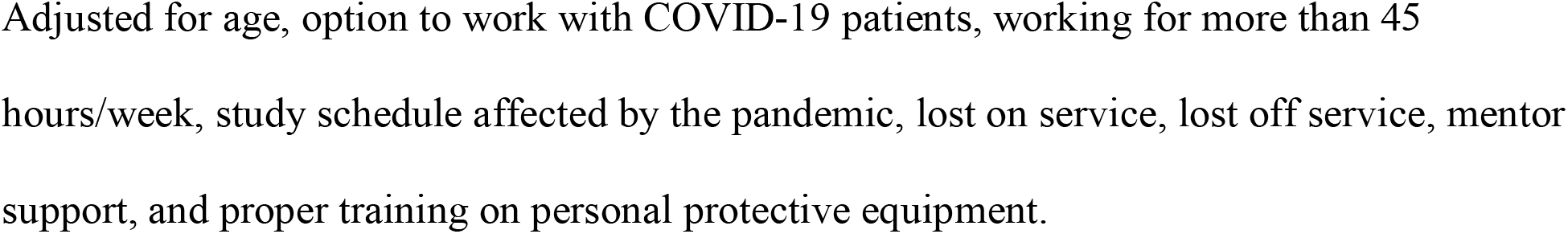
Regression analysis for depression

**Table 5.** Regression analysis for anxiety

| **Beta coefficient (95% confidence interval);P value** | **Beta coefficient (95% confidence interval);P value** |
| --- | --- |
| Male | Reference |
| Female | 3.1 (0.089-5.2);0.04 |
| Doctors feel that their training has been affected by the pandemic | 2.9 (0.04-5.9);0.047 |
| Lost on service | 2.5 (0.11-4.8);0.04 |
| Adjusted for gender, option to work with COVID-19 patients, working for more than 45 hours/week, lost on service, training affected by the pandemic, mentor support, and proper training on personal protective equipment. |  |

## Discussion

Our study indicated that various factors were independently associated with the mental state of residents. Gender, option to choose to work with COVID-19 patients, weekly working hours, training affected by the pandemic, lost on training service time were the factors associated with anxiety scores. While age, option to choose to work with COVID-19 patient, weekly working hours, working during the pandemic affected your study schedule, lost on and off service training time, support from your senior/supervisor/mentor, supervised training on personal protective equipment were associated with depression scores. The stress scores were associated with factors such as age, gender, option to choose to work with COVID-19 patient, weekly working hours, training, working during the pandemic affected your study schedule, support from your senior/supervisor/mentor, supervised training on personal protective equipment.

The median stress and anxiety scores were significantly higher in females as compared to males. The observed results are consistent with previous studies, such as the one done by Lai and co-workers (Lai et al., 2020). It is hypothesized that females experience higher stress than males because their stress responses are varied. Women have a hormonal system that is radically different from males, causing them to behave more emotionally. These distinctions between men and women occur during the reproductive years and gradually fade after menopause. Furthermore, they are confronted with more stressors since they are required to perform multiple roles in their daily activities (Verma et al., 2011).

In addition, the present study revealed that median scores of stress and depression were significantly higher among those aged 25-29 years and 29-33 years as compared to those aged 33-37 years. This could be related to the fact that most mental health problems begin in early adulthood, but young adults rarely receive mental health support. Furthermore, mental health problems are linked to a higher frequency of physical and emotional disorders in the medium to long term, marginalization, poor sleep quality, and dysfunctional interactions in the said population (Ramón-Arbués et al., 2020).

In our study, there was a strong association between longer working hours and higher DASS-21 score results. Long work hours have been linked to psychological and occupational stress in some research studies (Hsu et al., 2019; Lee et al., 2017). In addition, long working hours have been linked to an increased risk of cardiovascular disease, chronic fatigue, depressive state, anxiety, and self-perceived health, mental health status, hypertension, and health behavior (Liu et al., 2002; Artazcoz et al., 2009).

Varying types of job conditions are assumed to have different psychological effects due to disparities in the working environment, work intensity, burden, and disease risk. The major finding of the current study is that there is a clear link between high and extended work hours with the precipitation of depression, anxiety, and stress in healthcare workers during the pandemic era. The findings are consistent with the one reported by other researchers (Sasangohar et al. 2020; Lai et al. 2020). Because of the increased demand for healthcare services during the COVID-19 epidemic, many healthcare workers were forced to work harder, longer, or on more irregular schedules than they were used to do prior to the pandemic (WHO, 2020b). Longer working schedules in the assigned shifts, heavy workload and other psychosocial risks can lead to exhaustion, occupational burnout, elevated psychological discomfort, and negative mental health consequences (WHO, 2020b; Sasangohar et al., 2020). While it has been observed that professionals working in the intensive care unit in the battle against the pandemic are emotionally exhausted (Ruiz-Fernández et al., 2020). Teng et al. (2020) found that depression, anxiety, and fatigue are all frequent among frontline health workers.

A lack of supervised training on PPE, an insecure work environment, and poor working circumstances may lead to an elevated perception of risk to oneself and concern of transference to their family. This could lead to a significant decline of a drive as well as unpleasant emotions like despair and guilt. As a result, organizations need to ensure guaranteeing the safety of HCWs and addressing their basic requirements as a top priority. It’s also been discovered that peer support and supervisory assistance are linked to psychological well-being. The ability to converse to someone about their feelings or experiences, discuss the emotional and physical circumstances they face at work, and share their worries with other co-workers can all contribute to alleviatingfeelings of isolation and stress. Doctors on the job should be motivated to interact with one another, and if necessary, support groups should be accessible via social networks. Additionally, individual psychological load appears to be linked to sentiments of occupational expertise during COVID-19-related duties. Providing appropriate pre-job training for those who will take a job on the front lines, outlining precise and reliable information about the disease, the threat of contagion, and ability to treat oneself, as well as instituting systematic diagnostic and treatment procedures with specific guidelines, can enable alleviate stress and boost occupational esteem [Elbay et al., 2020]. Peer support programs have been shown to improve junior doctors’ psychological well-being and minimize the likelihood of burnout among clinicians (Chanchlani et al., 2018). A thriving trainee doctor peer support group will presumably foster a sense of connectedness and teamwork, as well as give substance to experiences, reducing burnout and improving patient care in the long run (West et al., 2018).

Real-life clinical situations, simulation, academic lectures, small group sessions, journal clubs, and teaching sessions are all used in the education program of the residents. However, the COVID-19 pandemic, on the other hand, has hampered the routine teaching and learning schedules of the residents. Considering medical education is so crucial to residents’ advancement in their careers, any disruption or alteration in their learning might have a negative impact on their success. During this time, online learning seemed to be the cornerstone of the learning process, since it could convey information while avoiding social contact, which could aid in virus spread. Many residents believe that their study routine was disrupted during the epidemic; this adjustment has been linked to increased levels of stress and depression in students, according to these research studies (Seifman et al., 2020).

Responders who perceived the pandemic had impacted their training and study schedule, who had lost service time, and who did not have the option to select whether or not to work with COVID-19 patients, or who were pushed to, had considerably higher stress levels. Internal medicine, emergency medicine, and family medicine residents were rerouted to higher-need areas. Some essential rotations, such as pediatrics and anesthesia, were canceled entirely in order for these redeployments to take place. In addition, due to a transition to digital care and efforts to curb resident exposure, chances to learn and enhance technical knowledge came to a standstill. This is in line with the findings of a reported study, which found that junior residents were more likely to possess more severe mental health consequences like depression, anxiety, insomnia, and discomfort (Lai et al., 2020). Lack of experience, insufficient preparation, or lack of training could all be plausible reasons for the observed effects. Healthcare executives hence need to create and implement support initiatives that are based on the needs and wishes of healthcare professionals. Peer counseling with connectivity to a therapist, therapy sessions and counseling, and an online clinician support group, for example, received the most interest in a survey (Shechter et al., 2020).

It is vital to assess the impact of limits in face-to-face work supervision, as well as the merits and implications of giving training through alternative methods such as online platforms and their possible permanent integration into the curriculum. Similarly, in light of trainee concerns about their capacity to meet specific training needs and pass competency assessments, competence definitions and curricula need to be re-evaluated (Seifman et al., 2021).

### Limitations of the study

The survey’s voluntary structure may have resulted in a selection bias, and the participants may not accurately reflect the actual population. A self-report questionnaire was employed to assess psychological symptoms, which did not concentrate on diagnostic evaluation by mental health specialists, in order to contact as many people as possible during this emergency situation and to reduce face-to-face interviews. We solely looked into the depression, anxiety, and stress levels of junior residents in this study. More research incorporating social support and post-traumatic stress disorder assessment among healthcare workers, on the other hand, would undoubtedly add to the literature.

## Conclusion

A pandemic necessitates swift organizational and healthcare personnel adaptability. It is therefore vital for organizations to consider factors that are independently associated with depression, anxiety, and stress which needs to be taken into consideration to protect the Residents’ Mental Well-being.

Supportive programs need to be envisaged for health professionals who are engaged in an adverse event. Organizations need to exploit the pandemic as a platform to devise focused initiatives to reduce significant stressors that affect healthcare employees’ mental well-being under normal conditions. The downturn has taken a tremendous toll on healthcare professionals.

Those in roles of institutional or sector leadership should seize this advantage to implement targeted initiatives to alleviate critical stressors that affect the mental health of healthcare professionals.

## Data Availability

All data produced in the present study are available upon reasonable request to the authors

## Acknowledgements

None

## References

Armocida, B., Formenti, S., Ussai, F., Palestra, F., & Missoni, E. (2020). The Italian health system and the COVID-19 challenge. Lancet Public Health, 5, e253. https://doi.org/10.1016/S2468-2667(20)30074-8

Artazcoz, L., Cortès, I., Escribà-Agüir, V., Cascant, L., & Villegas, R. (2009). Understanding the relationship of long working hours with health status and health-related behaviours. Journal of Epidemiology & Community Health, 63, 521–527. https://doi.org/10.1136/jech.2008.082123

Brunner, R. (2017). Why do people with mental distress have poor social outcomes? Four lessons from the capabilities approach. Soc Sci Med, 191, 160–7. https://doi.org/10.1016/j.socscimed.2017.09.016

Chanchlani, S., Chang, D., Ong, J.S., & Anwar, A. (2018). The value of peer mentoring for the psychosocial wellbeing of junior doctors: a randomised controlled study. Medical Journal of Australia, 209, 401–5. https://doi.org/10.5694/mja17.01106

Conceição, P., Hall, J., Jahic, A., Kovačević, M. S., Nayyar, S., Ortubia, A., & Tapia, H. (2020). COVID-19 and Human Development: Assessing the Crisis, Envisioning the Recovery. United Nations Development Programme. (Accessed 23 June 2021).

Coughlan, C., Nafde, C., Khodatars, S., Jeanes, A.L., Habib, S., Donaldson, E., Besi, C., & Kooner, G.K. (2021). COVID-19: lessons for junior doctors redeployed to critical care. Postgraduate Medical Journal, 97, 188–191. https://doi.org/10.1136/postgradmedj-2020-138100

Elbay, R.Y., Kurtulmuş, A., Arpacioğlu, S., & Karadere, E. (2020). Depression, anxiety, stress levels of physicians and associated factors in Covid-19 pandemics. Psychiatry Research, 290, 113130. https://doi.org/10.1016/j.psychres.2020.113130

Hsu, H.C. (2019). Age differences in work stress, exhaustion, well-being, and related factors from an ecological perspective. International Journal of Environmental Research and Public Health, 16, 50. https://doi.org/10.3390/ijerph16010050

Lai, J., Ma, S., Wang, Y., Cai, Z., Hu, J., Wei, N., & Tan, H. (2020). Factors associated with mental health outcomes among health care workers exposed to coronavirus disease 2019. JAMA Network Open, 3, 1–12. https://doi.org/10.1001/jamanetworkopen.2020.3976

Lee, K., Suh, C., Kim, J.E., & Park, J.O. (2017). The impact of long working hours on psychosocial stress response among white-collar workers. Industrial Health, 55, 46–53. https://doi.org/10.2486/indhealth.2015-0173

Liu, Y., & Tanaka, H. (2002). Overtime work, insufficient sleep, and risk of non-fatal acute myocardial infarction in Japanese men. Occupational and Environmental Medicine, 59, 447–451.

Lovibond, S.H., & Lovibond, P.F. (1995). Manual for the Depression Anxiety & Stress Scales. (2nd Ed.), Sydney: Psychology Foundation.

Raj, S.K. (2016). Well-being in residency: a systematic review. Journal of Graduate Medical Education, 8, 674–684. https://doi.org/10.4300/jgme-d-15-00764.1

Ramón-Arbués, E., Gea-Caballero, V., Granada-López, J. M., Juárez-Vela, R., Pellicer-García, B., & Antón-Solanas, I. (2020). The prevalence of depression, anxiety and stress and their associated factors in college students. International Journal of Environmental Research and Public Health, 17, 7001. https://doi.org/10.3390/ijerph17197001

RuizLJFernández, M. D., RamosLJPichardo, J. D., IbáñezLJMasero, O., CabreraLJTroya, J., CarmonaLJRega, M. I., & OrtegaLJGalán, Á.M. (2020). Compassion fatigue, burnout, compassion satisfaction and perceived stress in healthcare professionals during the COVIDLJ19 health crisis in Spain. Journal of Clinical Nursing, 29, 4321–4330. https://doi.org/10.1111/jocn.15469

Salem, J., Hawkins, L., Gates, J., Sundaram, A., Ong, Y.E., Fernando, A., Snelgrove, H., Mistry, M., Suleman, S. & Chakravorty, I. (2020). COVID-19 and the impact on doctor well-being and training-A mixed methods study. The Physician, 6. https://doi.org/10.38192/1.6.3.2

Sasangohar, F., Jones, S. L., Masud, F. N., Vahidy, F. S., & Kash, B. A. (2020). Provider burnout and fatigue during the COVID-19 pandemic: lessons learned from a high-volume intensive care unit. Anesthesia and Analgesia, 131, 106–111. https://doi.org/10.1213/ane.0000000000004866

Seifman, M. A., Fuzzard, S. K., To, H., & Nestel, D. (2021). COVID-19 impact on junior doctor education and training: a scoping review. Postgraduate Medical Journal. http://dx.doi.org/10.1136/postgradmedj-2020-139575

Shah, K., Kamrai, D., Mekala, H., Mann, B., Desai, K., & Patel, R. S. (2020). Focus on mental health during the coronavirus (COVID-19) pandemic: applying learnings from the past outbreaks. Cureus, 12(3), e7405. https://doi.org/10.7759/cureus.7405

Shechter, A., Diaz, F., Moise, N., Anstey, D. E., Ye, S., Agarwal, S., & Abdalla, M. (2020). Psychological distress, coping behaviors, and preferences for support among New York healthcare workers during the COVID-19 pandemic. General Hospital Psychiatry, 66, 1–8. https://doi.org/10.1016/j.genhosppsych.2020.06.007

Tang, F., Liang, J., Zhang, H., Kelifa, M. M., He, Q., & Wang, P. (2021). COVID-19 related depression and anxiety among quarantined respondents. Psychology & Health, 36, 164–178. https://doi.org/10.1080/08870446.2020.1782410

Teng Z, Wei Z, Qiu Y, Tan Y, Chen J, Tang H, Huang J (2020) Psychological status and fatigue of frontline staff two months after the COVID-19 pandemic outbreak in China: A cross-sectional study. J Affect Disord 275:247–252. https://doi.org/10.1016/j.bbi.2020.04.055

Torales, J., O’Higgins, M., Castaldelli-Maia, J. M., & Ventriglio, A. (2020). The outbreak of COVID-19 coronavirus and its impact on global mental health. International Journal of Social Psychiatry, 66, 317–320. https://doi.org/10.1177%2F0020764020915212

Verma, R., Balhara, Y. P. S., & Gupta, C. S. (2011). Gender differences in stress response: Role of developmental and biological determinants. Industrial Psychiatry Journal, 20, 4. https://dx.doi.org/10.4103%2F0972-6748.98407

West, C. P., Dyrbye, L. N., & Shanafelt, T. D. (2018). Physician burnout: contributors, consequences and solutions. Journal of Internal Medicine, 283, 516–529. https://doi.org/10.1111/joim.12752

World Health Organization. (2020). WHO calls for healthy, safe and decent working conditions for all health workers, amidst COVID-19 pandemic. World Day for Safety and Health at Work: WHO key facts & key messages to support the day. Available from: https://www.who.int/news-room/detail/28-04-2020-who-calls-for-healthy-safe-and-decent-working-conditions-for-all-health-workers-amidst-covid-19-pandemic

